# Acceptability and associated factors of indoor residual spraying for Malaria control by households in Luangwa district of Zambia: A multilevel analysis

**DOI:** 10.1101/2022.03.27.22272902

**Authors:** Maureen Aongola, Patrick Kaonga, Charles Michelo, Jessy Zgambo, Joseph Lupenga, Choolwe Jacobs

## Abstract

**Background:** The global burden of malaria has increased from 227 million cases in 2019 to 247 million cases in 2020. Indoor residual spraying (IRS) remains one of the most effective control strategies for malaria. The current study sought to measure the acceptability level and associated factors of indoor residual spraying.

**Methods:** A cross sectional study was conducted from October to November 2020 in sixteen urban and rural communities of Luangwa district using a cluster sampling method, Multilevel analysis was used to account for the hierarchical structure of the data.

**Results:** The acceptability level of indoor residual spraying among household heads was relatively high at 87%. Individuals who felt the timing was not appropriate were associated with decreased odds of accepting IRS (AOR = 0.55, 95% CI: 0.20 - 0.86). Positive attitude was associated with increased odds of accepting IRS (AOR = 29.34, 95% CI: 11.14 - 77.30). High acceptability level was associated with unemployment (AOR = 1.92, 95% CI: 1.07 - 3.44). There were no associations found between acceptability levels and community-level factors such as information, education, communication dissemination, awareness achieved through door-to-door sensitization, and public address system.

**Conclusion:** Acceptability level of indoor residual spraying was relatively high among households of Luangwa District suggesting that the interventions are more acceptable which is essential in reaching malaria elimination by 2030. Finding that community factors known to influence acceptability such as information, education and communication as well as awareness were not important to influencing acceptability suggests need for reinforcing messages related to indoor residual spraying and redefining the community sensitization approaches to make indoor residual spraying more acceptable.

## Introduction

Malaria still remains a global public health problem with an estimated 241 million malaria cases in 2020 in 85 malaria endemic countries increasing from 227 million in 2019, with most of this increase coming from countries in the WHO African Region.(1). Malaria is endemic in Zambia despite massive scale up of control efforts in the past decade and is one of the most important vector-borne infection of concern (2). Similar to most sub-Saharan African countries, indoor residual spraying (IRS) remains one of the strategies most effective for malaria control (3).

IRS is based on the principle that the sprayed insecticide leaves a residue of chemical on the interior wall of the house that is effective to kill mosquitoes resulting in the disruption of the disease transmission (4). However, acceptability by the households and community leaders is important to achieve high coverage (5,6) and accelerate the movement towards malaria elimination by the year 2030 (*7*).

Acceptability of IRS varies from country to country and is influenced by different factors. For example, household acceptability of IRS was 97.6% in Southern Mexico, 41% in Mozambique 29.4% in South Africa and 27.8% in eastern Ethiopia (8,9,10). WHO recommends that at least 80% of the households should be sprayed for the intervention to have an impact on transmission cycle. There are several factors that are associated with households’ refusal of IRS implementation. Among these factors are community understanding and beliefs about the purpose of an IRS program (8–10). Some members of the community have concerns on the negative effects of IRS and have fear of IRS program, which may lead to refusal of the intervention (10). Other concerns are in regard to spray residue discoloring inner walls, allergic reactions to the chemical, households being informed at short notice, challenges of furniture movement and not being available at the time of spraying (11–14). Furthermore, for malaria control strategies using IRS to be more effective, meaningful and sustainable, aspects such as community engagement, knowledge, attitudes and practices has to be taken into consideration (15).

Some areas in Zambia such as Luangwa district remain high malaria transmission zones despite several intervention strategies. According to the district malaria surveillance report for 2018, malaria incidence for Luangwa district was at 459 per 1000. In 2019, Luangwa district had 69.2% of sprayed houses after the implementation of IRS. The district had a gap of 10.8% to reach the minimum target of 80% for high-risk endemic areas (17). To our knowledge, limited evidence exists on acceptability of IRS by the households in Luangwa district, Zambia. Therefore, the current study set out to measure acceptability and associated factors of IRS for the control of malaria by the households in Luangwa district, Zambia.

### Implementation of IRS in Luangwa

Implementation of IRS in Zambia began in 2003, following the success of IRS by the private sector at the Konkola Copper Mines. Currently, the National Malaria Control Centre (NMCC) of the ministry of health is implementing IRS for malaria as part of an integrated vector management strategy. In 2010 the NMCC with President’s Malaria Initiatives support, expanded IRS to cover a total of 1.3 million structures in 54 districts, representing 75% of the districts in the country and protecting over 6 million people. From these structures Luangwa district targeted 4000 structures to be sprayed.

According to Malaria reports from Luangwa District Health Office it clearly stated that from the time IRS started, the operations were done once annually during the months of December and January. Government policy is that IRS is supposed to be carried out every six months because the chemical which was mainly used (Actellic) lasted for six months. A study revealed that the practice of conducting IRS once per year, compromises the effectiveness of the intervention (20). As a result, the district was still recording high malaria incidence rates in the country.

## Materials and Methods

### Study setting

The study was conducted in Luangwa district which is located in Eastern part of Lusaka Province from October to November 2020. Luangwa district is served by seventeen rural health facilities, two of which have both inpatient and outpatient services. Most households are primarily situated along the Luangwa River with fishing and small-scale agriculture being the primary livelihoods of the population. The geographical location of the district and given that most populations live along Luangwa River makes the district be more prone to having high prevalence of malaria cases especially with the marked seasonal patterns after the rains from December to April.

### Study design and population

A cross-sectional survey was conducted in November 2020 in Luangwa district to assess the acceptability of indoor residual spraying for malaria control. The cross-sectional study was chosen because it assesses the prevalence of disease in clinic based samples and can usually be conducted relatively faster and inexpensive (17). The study population was a subset of the target population who experienced indoor residual spraying for the past one year. Luangwa district has approximately 4672 number of households with a projected population of 35710 (18). The households were selected based on the eligibility criteria.

### Inclusion criteria

All households who were contacted and asked to spray their homes were included in the study

#### Exclusion criteria

All those who didn’t give consent to be included in the study and were not available at the time of data collection were excluded

### Sample size and sampling

This was a two-stage cluster sampling method and was on hierarchical structure. The hierarchy (multilevel) follows households as level-1, and communities as level-2 implying that households were nested in communities. The sample size was determined by the use of the prevalence formula with a proportion of 0.5, the sample size was 385, in order to adjust for the required sample size for cluster sampling a design effect (deff) of 2 was used which brought the sample size to 770 a 10 % non-response rate was adjusted and 856 households were invited for interview, 790 completed the interview yielding 92% response rate.

A probability proportional to size was used to determine the total number of households to be selected in each community. This was determined by dividing the total catchment population for a health facility over the total population for the district multiplied by the sample size. The study thus sampled 78 households from Luangwa Boma, 62 from Feira, 27 from Luangwa District Hospital, 61 from Mandombe, 16 from Mphuka, 19 from Jenairo, 25 from Kapoche, 18 from Kanemele, 65 from Luangwa high school, 70 from Katondwe mission, 77 from Chitope, 45 from Mangelengele, 70 from Kaunga, 39 from Kasinsa, 48 from Kavalamanja and 70 from Sinyawagora.

#### Data collection techniques and tools

Structured questionnaires were administered by trained research assistants and investigator to collect data at each of the sampled households using Magpi data collection tool. The questionnaire was all inclusive to gather information pertaining to community acceptability and associated factors involving interventions of indoor residual house spraying for malaria transmission.

## Variables

### Outcome variable

The outcome variable was acceptability which was defined as the extent to which people delivering or receiving a health care intervention consider IRS to be appropriate, based on anticipated or experienced cognitive and emotional response to the intervention (19). Acceptability was measured using a 5-point likert scale, with the following categories: strongly disagree, disagree, neutral, agree and strongly agree. Acceptability was determined by answering four questions on a scale of 0 to 4, with 0 indicating strongly disagree and 4 indicating strongly agree. The greatest possible score was obtained by multiplying the highest Likert scale code by the total number of questions. An individual was judged to have high acceptability if they had a score of 12 or higher, indicating at least 75% agreement with the implementation of the IRS (20).

### Independent variables

#### House hold level fixed effects

The household level factors included: timing of IRS, advised to wait before entering the house after spraying, employment status, residential, informed consent, attitude, gender and educational level.

### Community level random effects

The community level factors included: information, education and communication dissemination and conduct of awareness campaigns.

#### Data analysis and modelling

### Model diagnostics and adequacy checking

We used variance inflation factor (VIF) to check for multicollinearity among the independent variables with the cut-off point of 1/VIF not more than 0.1. None of the variables in the model suggested multicollinearity. The final model included possible interaction terms (household-level variables, community-level variables, and intra-level interactions) and none were significant. The statistical significance level was set at alpha 0.05 (two-tailed). All analyses were conducted with Stata 14.1 (Stata Corp., College Station, Texas, USA).

### Statistical analysis

We conducted descriptive statistics that included frequency analysis (percentages) for categorical variables. Univariable analysis was conducted to assess candidate variables as determinates for acceptability, was quantified by the OR and 95% confidence interval (CI). The effect of each independent variable on the outcome variable was assessed as candidate variable for the multivariable analysis using a significance level of p<0.2.

### Building multilevel model

In our study, we wanted to estimate the proportion variation due to chance of not accepting IRS rather than accepting it that lies between communities. Data was nested with two levels of hierarchy (individual households and communities). Household level one variables were nested within level two (communities). We constructed a null model (model without predictors) using the command: “xtmelogit” with an option of “var” and “or” to obtain odds ratios and the equation is

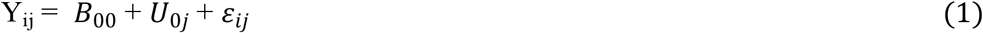

Where: Y_i*j*_ is the acceptability status of the i^th^ household in j^th^ community; *B*_00_is the probability of not accepting IRS without the independent variables; U_0*j*_ is the community-level effect and *ε*_i*j*_ is the error term at household level.

The intraclass correlation which represented the proportion of the between-community variation was calculated using the following formula:

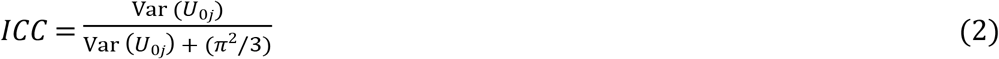

Where Var (U_0*j*_) is equal to the random intercept variance at the second level (community). As the value of Var (U_0*j*_) increases, the more the variation of odds between communities. The value of π^2^ (is approximately 3.29) from the standard logistic distribution. ICC represents the proportion of variation in not accepting IRS between communities which ranges from zero to one. If the ICC is closer to one (significant different from zero) suggests that there is significant variation in not accepting IRS between communities and if closer to zero (not significant different from zero) no variation and the traditional single level regression may be appropriate for analysis (21).

The null model showed that Var (U_0*j*_ of 4.91 and the ICC was 59% suggesting that multilevel regression model was appropriate than the traditional single level regression analysis. In this study 59% in variation in odds of not accepting IRS was explained by community-level differences.

Second, constrained intermediary model was constructed to determine the extent of variation in the household-level variables between communities. We estimated the variation of the effect of household variables on the odds of not accepting IRS from household to household to hospital since we thought this could be influenced by the services characteristics. A constrained was constructed with level-1 variables and community variables with no interaction between household-level variables with community-level variables(Aguinis, 2013). The following is the general equation:

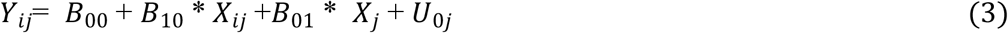

Where: X_i*j*_ is level-1 variables; X_*j*_ are level-2 variables (community variables); *B*_10_ is fixed slope for the household-level variables, and *B*_01_ is fixed slope for X_*j*_, which is the overall effect of the household level variables. This was followed by construction the augmented model which was done by testing household-level variables one by one. It is similar to the constrained model and the random slope variance suggests the variation of household-level variables from one community to the other and contains residual term associated with household-level variables. The following is the general equation:

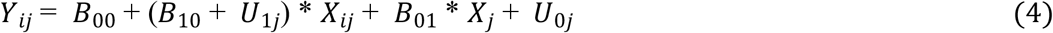

Where: U_1*j*_ represents the deviance of the community-specific slope which is the effect of household-level variables in a given community. The construction of the two models is to assess which one better fit to the data. Better fit was determined by likelihood ratio test and the smaller the deviance the better the fit.

Third, the final model was constructed to determine whether variables that can explain not accepting IRS and the following formula represents the model:

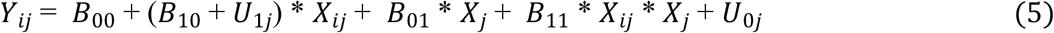

The equation is similar to equations two and three except that *B*_11_ estimates the coefficient for the interaction between household-level variables and community-level variables. In the analysis, the odds ratios were reported with the corresponding 95% confidence interval (CI).

### Interpretation of the final model

Before interpreting the final model, we considered a number of important aspects. First, Akaike Information Criteria (AIC) value for the random intercept was bigger compared to the one for random intercept and fixed coefficient model suggesting that the model with random intercept and fixed slope had a better fit than null model with random intercept in explaining not accepting IRS. Second, the deviance for the null model was bigger than the one for the random intercept and fixed slope model suggesting that the latter model was better (i.e., the lower the deviance the better the model). Third, the deviance-based chi-square value was significant suggesting that random coefficient model was better than single-level multiple logistic regression model in predicting not accepting IRS.

### Ethical clearance

This study obtained ethical clearance from University of Zambia Biomedical Research Ethics Committee (UNZABREC) (IRB 00001131) and National Health Research Authority of Zambia (000014/14/09/2020), Verbal informed consent was obtained from the participants before answering the questionnaire. All the participants had the chance to leave the study at any time.

## Results

Table 1 shows the social demographic and community characteristics of participants. Among the 790 participants, majority 353 (44.9%) were in the age range between 31 – 45 years, 442 (54.4%) were females and 566 (71.4%) were married. Most of the participants 492 (62.3%) had no or primary education and 487 (61.7%) were employed. More than three-quarters 696 (88.1%) were from rural areas and two-thirds 528 (66.8%) of the households had children under the age of 5 years. Almost everyone 787 (99.6%) indicated that they were given instructions for preparation of IRS, 769 (97.3%) were informed before spaying, 729 (92.3%) had positive attitude toward IRS and 783 (99.1%) were advised to wait after IRS. Majority 646 (81.8%) indicated that they had no side effects after IRS and 710 (89.9%) responded that the timing for IRS was no appropriate.

**Table 1:** Basic and community characteristics of household in Luangwa district.

| Basic characteristics |  | Community characteristics |  |
| --- | --- | --- | --- |
| Characteristic | Frequency (%) | characteristic | Frequency (%) |
| **Age (years)** |  | **Meeting held** | 62 (7.9) |
| < 30 | 192 (24.4%) | No | 728 (92.2) |
| 31- 45 | 353 (44.9%) | Yes |  |
| ≥ 46 | 242 (30.8%) |  |  |
| **Gender** |  | **Id card** |  |
| Male | 368 (46.6) | No | 765 (96.8) |
| Female | 422 (53.4) | Yes | 25 (3.2) |
| **Marital status** |  | **Community complies** |  |
| Single | 223 (28.3%) | No | 70 (8.9) |
| Married | 566 (71.7%) | Yes | 720 (91.1) |
| **Educational level** |  | **Awareness done** |  |
| No education or Primary | 492 (62.3%) | Meetings | 227 (28.7%) |
| Secondary | 244 (30.9%) | Door to door | 206 (26.1%) |
| Tertiary | 54 (6.8%) | PA system | 357 (45.2%) |
| **Employment status** |  | **IEC disseminated** |  |
| Employed | 487 (61.7%) | Meetings | 414 (52.4%) |
| Unemployed | 303 (38.4%) | Electronic print media | 376 (47.6) |
| **Residential** |  | **Age of operators (years)** |  |
| Urban | 94 (11.9) | ≤24 | 59 (7.5) |
| Rural | 696 (88.1) | 25-29 | 403 (51.0) |
|  |  | ≥ 30 | 328 (41.5) |
| **Number of children under 5 years** |  |  |  |
| None | 252 (31.9) |  |  |
| 1 – 3 | 528 (66.8) |  |  |
| ≥ 4 | 10 (1.3) |  |  |
| **Informed before spraying** |  |  |  |
| No | 21 (2.7) |  |  |
| Yes | 769 (97.3) |  |  |
| **Instruction for IRS** |  |  |  |
| No | 3 (0.4) |  |  |
| Yes | 787 (99.6) |  |  |
| **Timing** |  |  |  |
| timing not appropriate | 710 (89.9) |  |  |
| timing appropriate | 80 (10.1) |  |  |
| **Attitude** |  |  |  |
| negative | 61 (7.7) |  |  |
| positive | 729 (92.3) |  |  |
| **Side effects** |  |  |  |
| No | 646 (81.8) |  |  |
| Yes | 144 (18.2) |  |  |
| **Advised to wait** |  |  |  |
| No | 7 (0.9) |  |  |
| Yes | 783 (99.1) |  |  |

For community characteristics, majority 728 (92.2%) households participated in the preparatory meetings, 765 (96.8%) households revealed that spray operators had identity cards and most of the community participants 720 (91.1%) complied to IRS intervention. Most of the awareness campaigns 357 (45.2%) were mostly done through public address system. Slightly above half 414 (52.4%) had IEC disseminated through meetings and 403 (51.0%) households revealed that spray operators were in the age range between 25 – 29 years. All community members were aware, engaged and persuaded to participate in IRS programme.

## Acceptability Parameters

The questions that participants were asked regarding acceptability of IRS were on a 5-point Likert’s scale. In this study, the overall acceptability of IRS 687 (87%, [95% CI: 1.01-1.24]). More than half 442 (55.9%) of the participants agreed to the approach used, 442 (55.9%) agreed to the implementation approach, 428 (54.2%) liked the implementation and 422 (53.4%) welcomed the implementation of IRS (Table 2).

**Table 2:** Acceptability parameters and participants responses.

| Statement | Strongly disagree | Disagree | Neutral | Agree | Strongly agree |
| --- | --- | --- | --- | --- | --- |
| Does the approach used for IRS meets your approval? | 21 (2.7) | 7 (0.9) | 16 (2.0) | 442 (55.9) | 304 (38.5) |
| Is the implementation approach of IRS appealing? | 41 (5.2) | 19 (2.4) | 17 (2.2) | 442 (55.9) | 265 (33.5) |
| Do you like the implementation of IRS? | 29 (3.7) | 30 (3.8) | 17 (2.2) | 428 (54.2) | 286 (36.2) |
| Do you welcome the implementation of IRS? | 0 (0) | 10 (1.3) | 15 (1.9) | 422 (53.4) | 343 (43.4) |

### Multilevel predictors of indoor residual spraying acceptability

#### Final model

To arrive at the final model, we used the empty and intermediate model and being guided by the deviance of the augmented intermediated model and constrained model as well as the Akaike Information Criteria (AIC) the final model which is a combination of a random slope was found to be appropriate. Therefore, the results presented are for the adjusted multilevel analysis. In the univariate analysis to select potential variables for the adjusted multilevel analysis timing, side effects, attitude, advised to wait, informed about spraying and IEC dissemination were significant.

Table 3 shows the adjusted multilevel analysis, if accepted, production will need this reference to link the reader to the table. Household level factors timing not being appropriate for IRS significantly reduced the odds of accepting the intervention (OR 0.55, 95%CI: 0.20-0.86). Advised to wait before entering the household after IRS significantly increased the odds of accepting IRS by 8.47 times (95%CI: 1.00-72.13). Employment status increased the odds of accepting IRS (OR 1.92, 95%CI: 1.07-3.44) p value 0.029. Positive attitude among household heads increased the odds of accepting IRS (OR 29.34 95%CI: 11.14-77.30) p value 0.0001. The other variables were not significant.

**Table 3:**
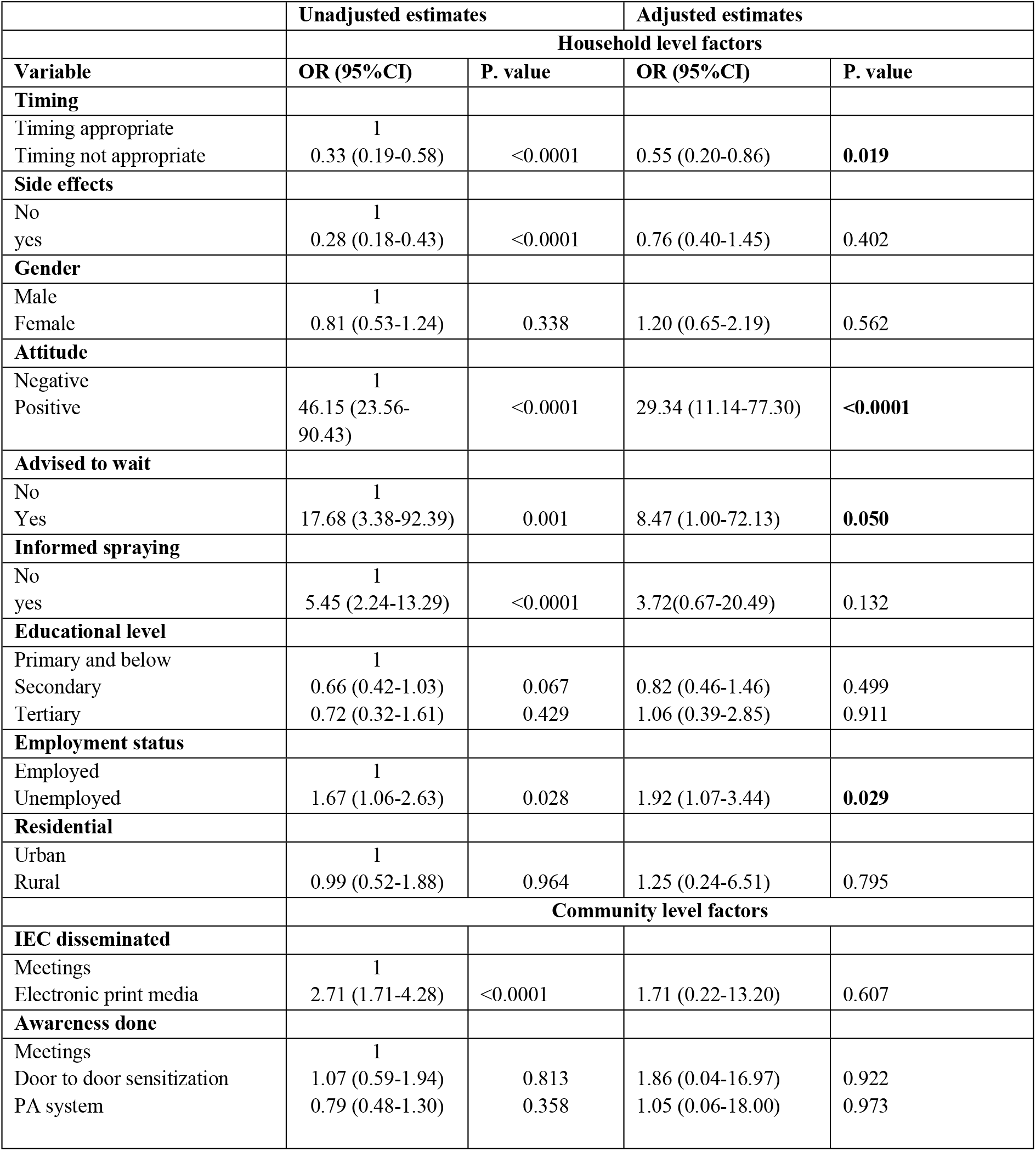
Multilevel analysis of predictors of IRS acceptability.

In the adjusted multilevel analysis, for community level factors Information, education and communication disseminated through electronic print media insignificantly increased the odds of accepting IRS intervention (OR 1.71, 95%CI: 0.22-13.20). Awareness of IRS campaign done through public address system insignificantly increased the odds of accepting IRS (OR 1.05, 955CI: 0.06-18.00) and the awareness done through door-to-door sensitization insignificantly increased the odds of accepting IRS intervention (OR 1.86, 95%CI: 0.04-16.97). Other known community level factors such as community awareness, community engagement and community member persuasion were not factors in this study because all the participants were aware, engaged and persuaded to participate in IRS programming.

## Discussion

This study was set to measure the level of acceptability of the interventions involving IRS among household’s heads in Luangwa District and factors contributing to the level of acceptability. The key findings of the study indicate that the overall acceptability of IRS among household heads was relatively high at 87%. The main factors in this current study that were found to contribute significantly towards the level of acceptability include timing, attitude, advised to wait and employment status. Although not significant, it was observed that community level factors such as dissemination of information, education and communication contributed to the level of acceptability.

In this study, we found relatively high variability in acceptability of indoor residual spraying across different settings. The acceptability level of IRS varies from one setting to another based on different influencing factors. In this current study the highest variability of acceptability was at 70% and lowest was 3%. This finding indicate that the difference may be attributed to the differences in the community settings and understanding of information education regarding indoor residual spraying. A study to identify community knowledge and acceptance of indoor residual spraying found that most participants who accepted the spraying were happy that the intervention was being done to combat malaria in their communities, but participants in some geographic areas felt it had limited effectiveness or safety. Additionally, participants from rural areas and whose homes had not previously been sprayed prior to their acceptance of the campaign were generally more satisfied and perceived IRS as more effective than those participants from urban areas (22). The implication of this finding is that combating malaria in these communities may remain a challenge. Therefore, there is need to reinforce information education on indoor residual spraying.

There are several household factors that contribute to acceptability of IRS, among the factors is timing of spraying, this current study revealed that timing of spraying has an impact on acceptance of IRS, these findings indicate that appropriate timing of spraying during the campaign can improve the acceptance of IRS intervention. A study on acceptability and perceived side effects of insecticide IRS found that timing of the spraying period is important in the implementation of IRS program and refusal rate to have the housing units sprayed, were high if the period is ill timed (28). Therefore, combating Malaria, based on IRS is associated with willingness of households to accept the spraying of residual insecticides during the spraying campaign (24).

Positive or good attitude towards IRS program is key in acceptance of IRS intervention (22). For example, a study in Uganda found that respondents with positive attitude were more likely to accept IRS intervention (25). Similarly, this study found that those with positive attitude had higher odds of acceptability than those with negative attitude. Reports suggests that attitudes and misconceptions related to acceptance or refusal of IRS found that knowledge and perception towards malaria lay a groundwork for acceptance or non-acceptance of malaria interventions ((26). Others have reported that prior IRS experience, impacts a negative attitude towards IRS as refusal was because they did not feel that the past campaigns were as effective as had been promised (32). However, certain behavior such as modification of the wall surface post IRS has potential to reduce actual IRS effectiveness and impact as fewer people are truly protected by IRS ((26).

Side effects associated with the chemicals used for IRS contribute to acceptance or refusal of the intervention, this study found that side effects had no significant impact on acceptance of IRS. These findings are contrary to a study which found that symptoms most frequently associated with a chemical used were headache, abdominal pain dysuria and vomiting. The most lasting symptoms were itching, sneezing and vomiting (33). A study found that a strong repugnant smell associated with IRS was reported to be a deterrent to IRS use. Participants reported developing allergic reactions including asthma and swelling of the face on entering a sprayed house (34). A study found that some householders were concerned about the health implications such as the exposure of asthmatic patients to the odor, and other householders associated the odor with inability to sleep (30).

On plausible explanation why there was a difference between this study and others could be due to small number in this study for those who indicated that they were not advised to wait (<1%) which could have affected statistical efficiency.

In the current study, community level factors such as information, education and communication dissemination to the households were not associated with acceptability. On a contrary, a study found that inadequate information and education prior to IRS increased the odds of non-acceptance (35).

This study has limitations that could have affected the findings of the study. First, assessment of acceptability was based on self-reported and may have affected the accuracy of measurements leading to “misclassification”. Participants were asked through a structured questionnaire and this has potential for desirability bias which we could not rule out. In that sense, we believe the reported estimates may be under or over estimated. Second, the inferent nature of a cross-sectional design could not support causal relationships. Despite the limitations our study brings out key issues around acceptability level of IRS. Also, the use of multilevel logistic regression analysis was appropriate considering clustering effect of the different communities to obtain reliable estimates and standard errors.

It is therefore, recommended that for future studies should utilize longitudinal design so that all variable can be observed and detect changes or developments in the study population over time. In this way accurate and causal relationship can be drawn.

## Conclusion

This study was set to measure acceptability levels and associated factors of IRS among household heads of Luangwa district. The findings suggest that acceptability level of IRS was relatively high proposing that the interventions are more acceptable among people of Luangwa district which is key in reaching malaria elimination by 2030. Although some factors such as community awareness, community engagement and persuasion of community members known to influence acceptability of IRS were not factors in this study, it will be important to determine what factors may have led to a high level of acceptability through further research. There is need for the district to improve on the starting time of IRS if the desired impact of preventing malaria is to be achieved.

## Data Availability

no legal or ethical resyrictions

## Acknowledgements

We thank the data collection team from Luangwa District health facilities spearheaded by Edson Musonda for their assistance and support in conducting interviews. We also thank Professor Joseph Mumba Zulu for his contributions during manuscript development. Additionally, the corresponding author is a recipient of a TDR scholarship under the postgraduate training scheme in implementation research at the University of Zambia, School of Public Health. We are grateful for the support from the training scheme, as provided by the UNICEF/UNDP/World Bank/WHO special programme for Research and Training in Tropical Diseases (TDR).

## Author Contributions

MA designed the study, coordinated recruitment of participants, collected & entered data, analyzed & drafted the manuscript. CJ designed the study, participated in data analysis & reviewed draft manuscript. PK guided data analysis & reviewed draft manuscript. JZ, JL, CM designed the study & reviewed draft manuscript. All authors read & approved the final manuscript.

## Notes

### Competing Interest Statement

The authors have declared no competing interest.

### Funding Statement

No source of funding

### Author Declarations

The study obtained ethical clearance from the University of Zambia Biomedical Research Ethics Committee (UNZABREC) (IRB00001131) and the National Health Research Authority of Zambia (000014/14/09/2020)

